# Interplay between polygenic effects and polypharmacy on dementia: An investigation in an elderly Scottish cohort

**DOI:** 10.1101/2024.11.01.24316584

**Authors:** Vasilis Raptis, Donncha Mullin, Sumbul Syed, Ian J Deary, Simon R Cox, Tom C Russ, Michelle Luciano

## Abstract

**INTRODUCTION:** Polygenic Risk Scores for Alzheimer dementia (AD-PRS), a measure of aggregate AD genetic risk and polypharmacy have been associated with dementia. Here, we test their interaction’s association with future dementia among older adults without baseline neurodegenerative diagnoses.

**METHODS:** Using Cox proportional hazards and mortality-adjusted competing risk regression models we analysed up to 17.5 years all-cause incident dementia in the Lothian Birth Cohort 1936 (n=759, 105 dementia patients). We used polypharmacy (total or nervous-system-acting medications count), AD-PRS, and their interaction as main predictors.

**RESULTS:** A non-significant interaction was found between AD-PRS and total polypharmacy (HR=1.06; p=0.15) or nervous-system-acting polypharmacy (HR=0.98; p=0.86) in shaping dementia risk. Omitting interaction, mortality-adjusted models showed significant AD-PRS prediction of dementia (HR ∼1.40; p<0.001), non-significant total (HR=1.03; p=0.49), and nervous-system-acting polypharmacy effects (HR=1.27; p=0.069).

**DISCUSSION:** Elucidating the complex interplay between polypharmacy and genetics could improve management of inappropriate medication in older adults genetically prone to dementia/AD.

## 1 BACKGROUND

Dementia, a condition describing the significant and progressive overall deterioration of a person’s cognition with consequent deterioration in function^1^, is widely recognised to have multi-factorial causes, arising from the interplay between environmental, lifestyle and genetic factors^2,3^.

Alzheimer dementia (AD), the most frequent cause of dementia (accounting for 70 - 80% of dementia^4^) has been extensively shown to have a strong genetic component^5–7^. Multiple genetic variants have been identified as risk factors for AD through large-scale Genome-Wide Association Studies (GWAS)^5,6^. The most unequivocally established AD risk gene is apolipoprotein E (*APOE*), located on chromosome 19 ^8–11^. Its coding protein, Apo-E, is an important cholesterol carrying protein, implicated in several brain functions, including lipid transfer, injury repair, neuroinflammation and others. Notably, the *APOE* ε4 allele confers a dramatic increase in AD prevalence (4 to 10 fold increase) and lower age of onset^8^, thus being considered an important genetic risk factor for AD.

Other smaller effect AD-associated genes have been implicated in multiple biological processes, including microglia involvement, tau and amyloid-β protein metabolism regulation, immunity, inflammation, cholesterol metabolism, and neurotransmitter regulation^12,13^. To study the contribution of many genetic loci with small effect, Polygenic Risk Scores (PRS) are often implemented, which estimate a single aggregate score for each individual that is indicative of their genetic liability to a disease^14^. For AD, PRS calculated using many AD associated genes have been found in several studies, with varying accuracy, to be predictive of AD risk, even without including the stronger *APOE* genetic effect^5,6,10,11^. Furthermore, AD-PRS have been associated with cognitive impairment in older healthy adults, as well as several AD related neurodegeneration phenotypes (e.g. neuroimaging changes)^12^.

Although individuals’ genetic variation is fixed at birth, genetic effects are thought to be modified through their interactions with environmental, lifestyle or social factors throughout life. Such gene by environment (GxE) interactions have been suggested in dementia and AD^3,15^. Specifically for GxE interactions in dementia, it has been proposed that genetic risk for dementia can be mitigated by favourable modifiable lifestyle factors^16–18^ (i.e., healthy diet, physical activity, non-smoking, and low alcohol consumption), although with sometimes conflicting results^17^. On the other hand, two randomised clinical trials^19,20^ assessing the effect of multidomain lifestyle interventions on dementia found that *APOE* ε4 allele carriers and non-carriers had similar results, indicating no *APOE* ε4 interplay with lifestyle factors. However, most studies so far have focused only on *APOE* genetic variants for GxE interactions^16,18–20^.

Regarding AD-PRS interaction with the environment / lifestyle in shaping dementia or AD risk, less work has been done. In a Chinese cohort study^15^, individuals living around higher residential greenness had lower chances of cognitive impairment, with the effect being increased in those having lower PRS genetic risk, although marginally significant. In another study, conducted on a large European cohort, adherence to a healthy lifestyle did not significantly interact with AD PRS in shaping dementia risk^2^.

Polypharmacy, a term indicating that someone takes multiple medications, is highly prevalent in older adults with multiple chronic conditions^21,22^. It has been demonstrated that polypharmacy is associated with unfavourable outcomes, such as increased frequency of falls, increased hospitalisation and hospital readmission, adverse drug reactions and increased mortality^21,23^. Harmful polypharmacy effects may arise due to drug-drug and drug-disease interactions, especially in older people who have a reduced ability to metabolise drugs^24^. Specifically for dementia, the role of polypharmacy as a risk factor is less known^25^. It has been estimated that polypharmacy burden is higher than the general population, given the higher number of comorbid physical conditions in dementia patients^22,26^, and polypharmacy in people with dementia has been associated with increased emergency hospitalisation and mortality^27^.

Whether the effect of polypharmacy on dementia risk can be modulated by genetic factors is not known. In the present study, it is hypothesised that individuals exposed to polypharmacy have an increased dementia risk if they also have a high genetic risk profile, compared to individuals with a low genetic risk profile. This hypothesis is based on the idea that biological pathways which may be disrupted by drug-disease and drug-drug interactions are also affected by the genetic variants that lead to dementia pathology. For example, anticholinergic drugs, acting on acetylcholine neurotransmitters, have been found to increase dementia risk, adverse outcomes, and are frequent in dementia patients with polypharmacy^22,28^. At the same time, acetylcholine neurotransmission-related genes (e.g. *RAB10* gene) have been linked with AD pathogenesis^13^. In this study, the proposed hypothesis is addressed in a longitudinal Scottish cohort of older adults, who were relatively healthy and dementia-free at the baseline age of 70 years. Specifically, an interaction effect between Alzheimer Disease Polygenic Risk Scores and the number of (1) total and (2) nervous system active baseline drugs is tested in terms of predicting dementia up to 17.5 years of follow-up.

## 2 METHODS

### 2.1 Dataset description

The present analysis was conducted on the Lothian Birth Cohort 1936 (LBC1936) dataset^29^. This is a longitudinal study, designed as a follow-up to the Scottish Mental Survey of 1947 (SMS1947), an intelligence test conducted on pupils aged 11 years old across Scotland^30^. From 2004 to 2007, LBC1936 recruited 1,091 participants of the SMS1947, then aged around 70 years old, and mostly living in Lothian at that time^29^. Since it focussed on ‘normal’ cognitive ageing initially, no participants had a diagnosis of dementia at baseline. After this first wave of recruitment, participants were re-invited to the study every three years (at mean ages: 73, 76, 79 and 82 years) for follow-up testing, termed Waves 1 to 5 respectively. Wave 6 has recently finished, but data is not yet available. At each wave, a wealth of data has been collected from questionnaires, blood tests, physical, medical and cognitive measurements. In addition, participants who have consented to do so, have been linked to their electronic health record to obtain further information about their health status (e.g., disease diagnoses, hospital visits). Detailed descriptions of the LBC1936 have been previously reported^29,31,32^. For this analysis, information from Wave 1 has been utilised, regarding: participants’ demographic characteristics, genetic data and self-reported medication usage and medical conditions. The analysis was conducted on 759 individuals (105 dementia patients, 654 controls) for whom complete genetic data (including *APOE* gene alleles) and ascertained dementia outcome as of August 2022 (see “Outcome: incident dementia” section) were available.

### 2.2 LBC1936 genetic data

During Wave 1 of the LBC1936, genome-wide genotyping and genotyping of the Apolipoprotein E variants (ε2, ε3, and ε4 alleles) was performed^29,32^ for 1,005 individuals. Genotyping of approximately 500,000 genetic variants across the genome (Single Nucleotide Polymorphisms, SNPs) was done using the Illumina Human610-Quadv1 chip^33^. Furthermore, non-genotyped variants have been imputed based on the Haplotype Reference Consortium^34^ reference panel, totalling 39 million SNPs.

High quality SNPs and individuals were extracted, in order to minimise genotyping and imputation errors and reduce the chance of biased results^35^. The quality control filters applied on genotyped data were described in Houlihan et al, 2010^33^.I In brief theyincluded the following: One of each related pair of individuals (> 2^nd^ degree relatives, i.e., half-siblings) were excluded (n=8); 1 individual with mostly non-European descent was excluded; 12 individuals whose reported and genetically inferred sex did not match (likely indicative of errors during genotype sample preparation) were excluded, and; SNPs that were missing from more than 2% of the individuals were excluded. For the present analysis, and on the genotyped and imputed data: 4,360,232 SNPs whose alleles were found in a frequency less than 1% in the cohort were excluded; 55 SNPs that deviated significantly from Hardy-Weinberg Equilibrium were excluded, and; 27.7 million imputed variants with an imputation score less than 0.8 were excluded. These quality control filtering steps were performed using the PLINK(v1.9) software^36,37^.

799 individuals remained after quality control, containing approximately 7 million SNPs. 759 individuals formed the final sample for the analysis, after individuals with missing values in the *APOE* predictor and the covariates were removed (see below).

### 2.3 Predictors

#### 2.3.1 Polygenic risk scores

Polygenic Risk Scores (PRS) were used as a risk factor representing the aggregated genome-wide genetic effect contributing to AD. PRS are calculated for each individual as a sum of their genome-wide risk alleles, weighted by each allele’s effect size to the outcome (here AD status)^14^. For AD-PRS calculation, the allele effect sizes were obtained from the largest to-date European GWAS on AD, totalling up to 85,934 AD patients (Bellenguez et al, 2022)^5^. AD was chosen, as this is the dementia sub-type for which the largest GWAS has been conducted, and the most frequent dementia sub-type in the general population and the present sample (48.6%, Supplementary Table 1).

The PRSice2 (v2.3.5) software^38^ was used to calculate the AD-PRS. PRSice2 uses the “clumping and thresholding” technique^14^. This means that the total number of SNPs is thinned down to retain only SNPs that are independent from each other (clumping) and significantly associated with the outcome (thresholding). Furthermore, the *APOE* region (chromosome 19, base pair 44,000,000 to 46,500,000^11^, in the human reference genome GRCh37 coordinates) was removed before the calculation of the AD-PRS (see “*APOE* ε4 alleles” section). Finally, AD-PRS were calculated for the 759 participants with non-missing data, using 981 genome-wide AD associated SNPs at a p-value threshold of < 0.0002.

#### 2.3.2 *APOE* ε4 alleles

Given the strong effect of the *APOE* ε4 polymorphisms on dementia, it has been suggested for genomic analyses to treat *APOE* polymorphisms independently as a separate predictor^10,11^, as it can mask the effects of other genetic variants with weaker effects. The *APOE* ε4 polymorphism is thus included as a separate risk factor in the present analysis, defined as the number of ε4 alleles per individual (0, 1 or 2). Since 32 individuals did not have information on *APOE* status, they have been excluded from the analysis, as this is an important risk factor for dementia.

#### 2.3.3 Polypharmacy

Polypharmacy was defined as the count of total or nervous system-acting drugs taken at baseline. Measurements were obtained using LBC1936 participants’ medication data. For each wave of the LBC1936, medication data was based on participants’ prescription cards which were brought to the assessment centre and further inspected by the testing team. Data includes prescribed drugs (in their brand name or their generic name), over-the-counter drugs and dietary supplements. To ensure consistency, (e.g., no spelling mistakes, multiple brand names referring to the same drug), we applied several pre-processing steps in order to code each drug name according to the Anatomical Therapeutic Chemical (ATC) classification system^39^. The ATC classification system classifies the active ingredient(s) of a drug in a 5-level hierarchical code, based on its acting organ/anatomical system and its therapeutic, pharmacological, or chemical properties.

Specifically, using only medication data from Wave 1 of LBC1936, the pre-processing steps included:

- Drugs in their brand names were searched in the British National Formulary (BNF) (https://bnf.nice.org.uk/)^40^ and substituted with their generic name.
- Drugs in their generic name were inspected and corrected for spelling mistakes, also using the BNF website.
- Subsequently, for each drug’s generic name, its corresponding ATC code was retrieved from the ATC classification system’s online searchable database (https://www.whocc.no/atc_ddd_index/)^39^. For drugs with multiple ATC codes (e.g., acting on more than one system), all available ATC codes were retrieved.
- The following entries were excluded from the analysis:

- Drugs that could not be assigned to an ATC code
- homeopathic substances
- diet supplements
- vitamins.
- Additionally, it was deemed appropriate to exclude topically applied dermatological drugs and those with ophthalmic, otic or nasal routes of administration, as they are unlikely to have a systemic effect and interact with pathways affected by AD related genes.

After these pre-processing steps were applied, a “general” polypharmacy variable was defined as the total number of drugs taken at Wave 1. Additionally, a nervous-system-specific polypharmacy variable was defined as the number of drugs acting on the central nervous system, based on the corresponding ATC code (code starting with “N0” ^39^).

#### 2.3.4 Other covariates

Apart from the PRS, *APOE* and polypharmacy variables, the following variables were included as covariates: sex, age, BMI, Scottish Index of Multiple Deprivation (SIMD) index, smoking status (non-, ex-, or current smoker), alcohol consumption status (yes or no), number of comorbidities (ma× 10). The number of comorbidities was calculated by summing the presence, at the time of Wave 1 assessment, of self-reported diseases among: high blood pressure, diabetes, high cholesterol, history of cardiovascular disease, stroke, neoplasia, arthritis, Parkinson’s disease, thyroid disease, leg pain or other unspecified disease. Dates of death were identified via data linkage with the National Health Service Central Register, provided by National Records of Scotland. Participant deaths were flagged to the research team approximately every 12 weeks beginning at study baseline. Finally, to account for any residual genomic population stratification^41^, which could inflate the effect of the PRS, the 4 first genomic principal components of the individuals were included as covariates.

### 2.4 Outcome: incident dementia

In a recent endeavour (as described here^42,43^), dementia outcomes have been ascertained for LBC1936 participants. Briefly, a multi-disciplinary team of clinical dementia experts provided consensus status for dementia and, when possible, dementia sub-types, for all consenting participants^42^. This process included a comprehensive review of participants’ electronic health records and death certificates for the presence of dementia. Additionally, when deemed necessary, home visits to suspected dementia patients were conducted by a trained clinician. Finally, a consensus diagnostic review board of experts assessed the strength of evidence for each participant, and categorised each as having “probable”, “possible”, or “no” dementia, as well as each dementia subtype. The dementia and dementia subtype diagnoses were based on the International Classification of Diseases-11 criteria^42,44^. Furthermore, the age of onset has been reported as the earliest age for which any diagnosis was reported. Age of onset spanned a time period from approximately one year after attending Wave 1 attendance until mid-August 2022, when the ascertainment process ended^42^.

In the present analysis, only “probable” dementia diagnoses were considered, due to the lack of definite evidence for the assignment of a dementia diagnosis in participants with “possible” dementia^42^. The outcome used here was time-to-onset, the time period between Wave 1 assessment until all cause (non-subtype specific) dementia diagnosis in 105 participants.

### 2.5 Statistical Analysis

The aim of the present study was to determine the effect of AD-PRS, baseline polypharmacy, and their interaction on incident dementia over up to 17.5 years of follow-up. Given the longitudinal nature of the outcome, Cox proportional hazards (Cox PH) models^45^ were implemented. This type of modelling investigates the association between multiple predictors and the time that passes until an outcome event occurs^46^ (here dementia).

In a further analysis, to account for the possible bias of individuals dying before exhibiting the main dementia outcome, the Fine-Gray competing risk model^47,48^ was used. This extension of the Cox proportional hazards model estimates the effect of a predictor on the outcome of interest (dementia), given that a competing event has not happened yet (death)^46^. Mirroring the procedure from Mullin et al. (2023)^43^, for both analyses the time-to-event outcome variables were calculated as the time passing between Wave 1 assessment and the earliest of the following: (1) dementia diagnosis, if the individual was ascertained for dementia, (2) the individual’s death, if the participant had died while dementia-free, or (3) end of ascertainment, if the individual was dementia-free and alive at the end of the ascertainment period (mid-August 2022). Statistical models were implemented using the predictors described previously (“Predictors” section), as well as the interaction between AD-PRS and the polypharmacy variables. The results of the models are in the form of Hazard Ratios (HR) with 95% confidence intervals (CI) for each tested predictor.

A null model that included only PRS and polypharmacy main effects, without their interaction was also implemented. The statistical analysis for those models was the same as described above.

As a sensitivity analysis, cox PH and competing risk models were also run using time to AD diagnosis as outcome (n=51), as described above for the all-cause dementia models.

All analyses were conducted using the R (v4.2.2) statistical software^49^, and the packages “survival”^50^ for Cox PH models and “cmprsk”^48^ for competing risk models.

## 3 RESULTS

### 3.1 Sample characteristics

After genotyping data cleaning and exclusion of individuals with missing values, the analysis sample consisted of 759 individuals. 105 individuals (13.8%) were diagnosed with dementia up to 17.5 years after follow-up from Wave 1. 51 of these individuals (6.7%) had an Alzheimer dementia diagnosis. Participants’ characteristics at Wave 1 for all tested covariates are presented in Table 1. Participants’ mean age at Wave 1 was 69.5 (standard deviation=0.83) years, while the mean age-at-onset for dementia was 80.7 (3.7) years. The median time from baseline to dementia diagnosis for later patients was 11.8 years [interquartile range (IQR): 7.88-14.35]. 277 (49.5%) participants had passed away at the time of censoring,

**Table 1:**
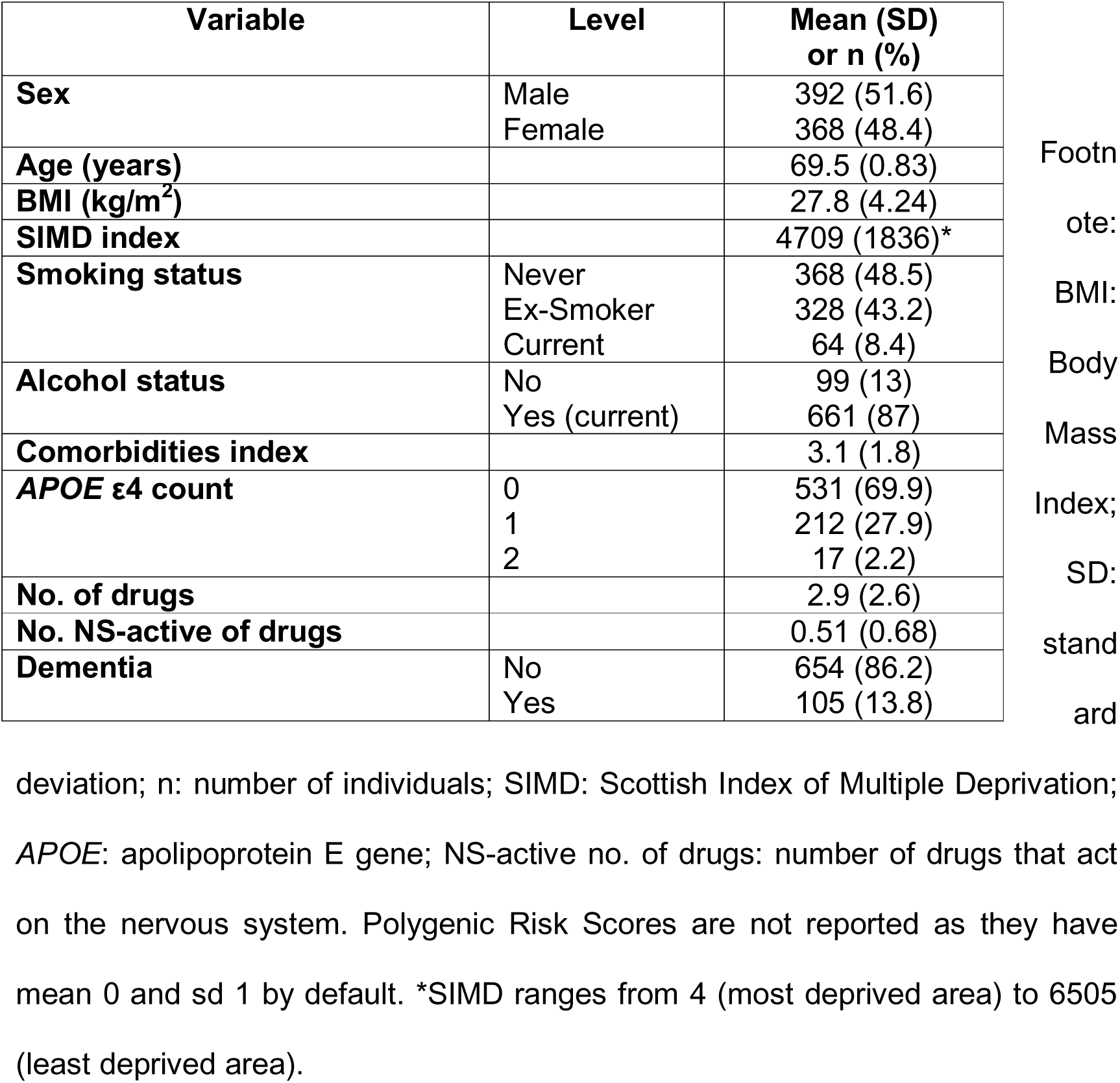
Descriptive statistics of LBC1936 at Wave 1.

Regarding polypharmacy, 164 (21.6%) of participants took 5 to 10 drugs at the time of Wave 1 testing, while 14 (1.8%) took more than 10 drugs simultaneously (excessive polypharmacy) (Sup. Table 2 and Sup. Figure 3), with a maximum of 16 simultaneous drugs. The most abundant drugs by the anatomical system on which they act (according to the ATC classification system^39^), were those acting on the gastrointestinal system / metabolism (346 individuals taking at least 1, 45.6%), cardiovascular system (388 individuals, 5.1%) and nervous system (321 individuals, 42.3%) (Sup. Table 2). In addition, individuals taking multiple medications acting on the same system are reported: 141 (18.6%) individuals taking 3 or more cardiovascular medications; 46 (6 %) individuals taking 3 or more gastrointestinal / metabolic medication; and 9 (1.2%) individuals with 3 or more nervous system active medications simultaneously (Table 1 and Sup. Table 2).

### 3.2 Main analysis

The Cox PH models (unadjusted for death as competing risk) were run using dementia status as the time-to-event outcome variable and polypharmacy (number of drugs), Polygenic Risk Score (PRS) and their interaction (polypharmacy*PRS) as predictors. The models were adjusted for the covariates: *APOE* ε4 count, age, sex, BMI, SIMD index, comorbidities index, alcohol status, smoking status and 4 first genomic principal components. The polypharmacy predictor was either the “general” polypharmacy (total number of drugs) or the nervous system active polypharmacy (number of drugs acting on the nervous system). Cox PH models utilising, among others, age at baseline, sex, comorbidities index and socioeconomic variable as predictors of dementia in LBC1936 have been reported by Mullin et al (2023)^43^. However, in this previous study motoric cognitive risk instead of polypharmacy / AD-PRS was the main predictor and Wave 3 was the baseline. Both here and in Mullin et al (2023)^43^ these covariates had a non-significant effect on dementia risk.

The results for the “general” polypharmacy Cox PH model are presented in the Figure 1 forest plot. Only the *APOE* ε4 allele count significantly increased – more than tripled – the risk of incident dementia over the 17.5-year follow-up (HR = 3.28; 95% CI: 2.38 – 4.54; p = 5.35×10^−13^). Taking more drugs and having a high PRS both increased the risk of incident dementia, however, the effects were not significant. For number of drugs: HR = 1.04; 95% CI: 0.93 – 1.15; p = 0.517, and for PRS: HR = 1.15; 95% CI: 0.85 - 1.55; p = 0.376. The interaction effect between the number of drugs and PRS was in the direction of increased dementia risk above that of their separate effects but was non-significant (HR = 1.06; 95% CI: 0.98 - 1.15; p = 0.135).

**Figure 1:**
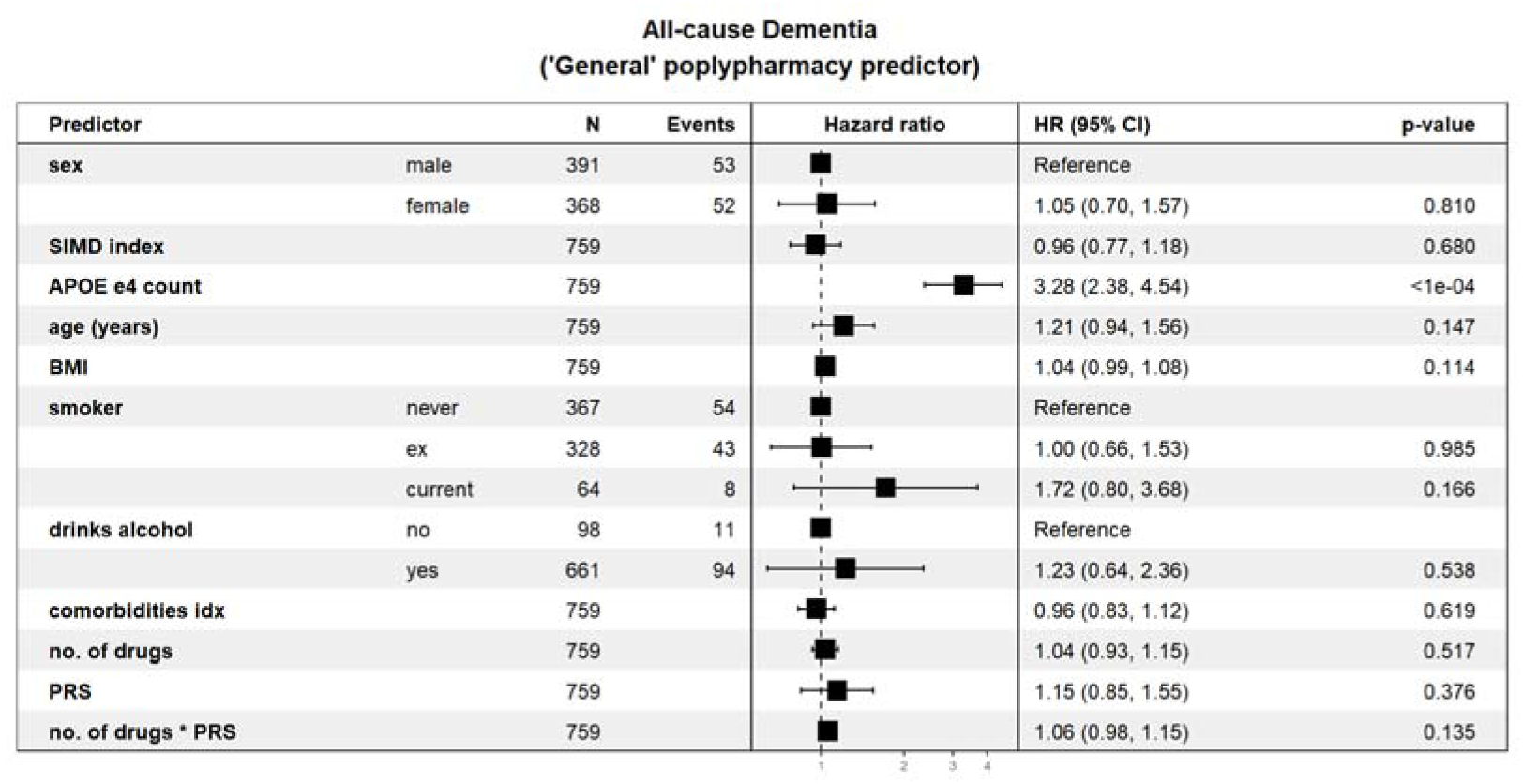
Forest plot showing the Cox PH model Hazard Ratios for each predictor’s association with all-cause incident dementia. The “no. of drugs” predictor includes all drugs taken by an individual after data cleaning. HR for Genomic Principal Components are not shown for illustration purposes, but were included in the model. The forest plot was produced using the “forestmodel” R package. HR: Hazard Ratio; CI: Confidence Interval; PRS: Polygenic Risk Score; BMI: Body Mass Index; SIMD: Scottish Index of Multiple Deprivation.

Figure 2 presents the association results for the Cox PH model that includes the “nervous system active” polypharmacy predictor. The *APOE* ε4 predictor is, again, a highly significant risk factor (HR = 3.36; 95% CI: 2.43 – 4.65); p = 2.63×10^−13^). This time, however, the number of nervous system active drugs significantly increased the follow-up dementia risk (HR = 1.37; 95% CI: 1.02 – 1.84; p = 0.035). The positive effect of the PRS was, now, also significant (HR = 1.42; 95% CI: 1.10 – 1.82; p = 0.006). The interaction between the number of nervous system active drugs and the PRS, however, had a non-significant negative effect (HR = 0.95; 95% CI: 0.72 – 1.25; p = 0.707).

**Figure 2:**
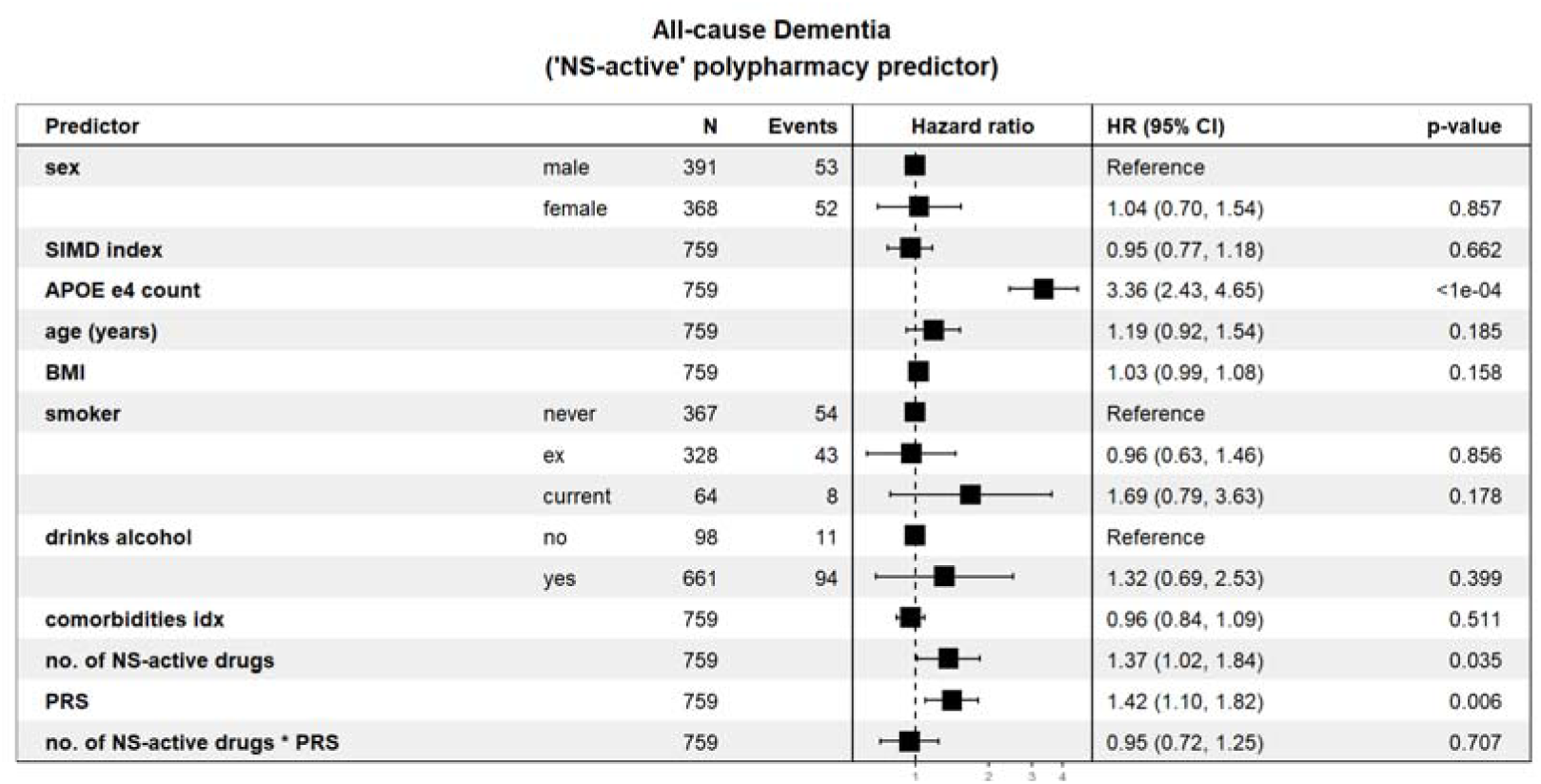
Forest plot showing the Cox PH model Hazard Ratios for each predictor’s association with all-cause incident dementia. The “no. of NS-active drugs” predictor includes drugs acting on the nervous system taken by an individual. HR for Genomic Principal Components are not shown for illustration purposes, but were included in the model. The forest plot was produced using the “forestmodel” R package. HR: Hazard Ratio; CI: Confidence Interval; PRS: Polygenic Risk Score; BMI: Body Mass Index; SIMD: Scottish Index of Multiple Deprivation; NS-active: nervous system-active.

The main results of the Fine – Grey competing risk regression models, adjusting for mortality as a competing risk of dementia, are presented in Figure 3. Models were run for both the “general” polypharmacy predictor and the “nervous system-active” polypharmacy, and their interaction with PRS, adjusting for the full set of covariates. Results for the full list of covariates are presented in Sup. Figure 4. For the “general” polypharmacy model, the results were similar to the unadjusted model, with only the *APOE* ε4 predictor having a significant effect (HR = 3.38; 95% CI: 2.46 – 4.66; p = 8.0×10^−14^). “General” polypharmacy, PRS and their interaction all have small, positive, non-significant association with incident dementia, after adjusting for the risk of death (Figure 3, top). Regarding the “nervous system-active” polypharmacy model, the PRS genetic effect remained significant after accounting for death (HR = 1.42; 95% CI: 1.13 – 1.78; p = 0.0029), but the “nervous system-active” polypharmacy predictor marginally lost significance (HR = 1.27; 95% CI: 0.98 – 1.65; p = 0.068). The PRS * “ns-active” polypharmacy interaction was again nonsignificant (HR = 0.98; 95% CI: 0.78 – 1.23; p = 0.86) (Figure 3, bottom).

**Figure 3:**
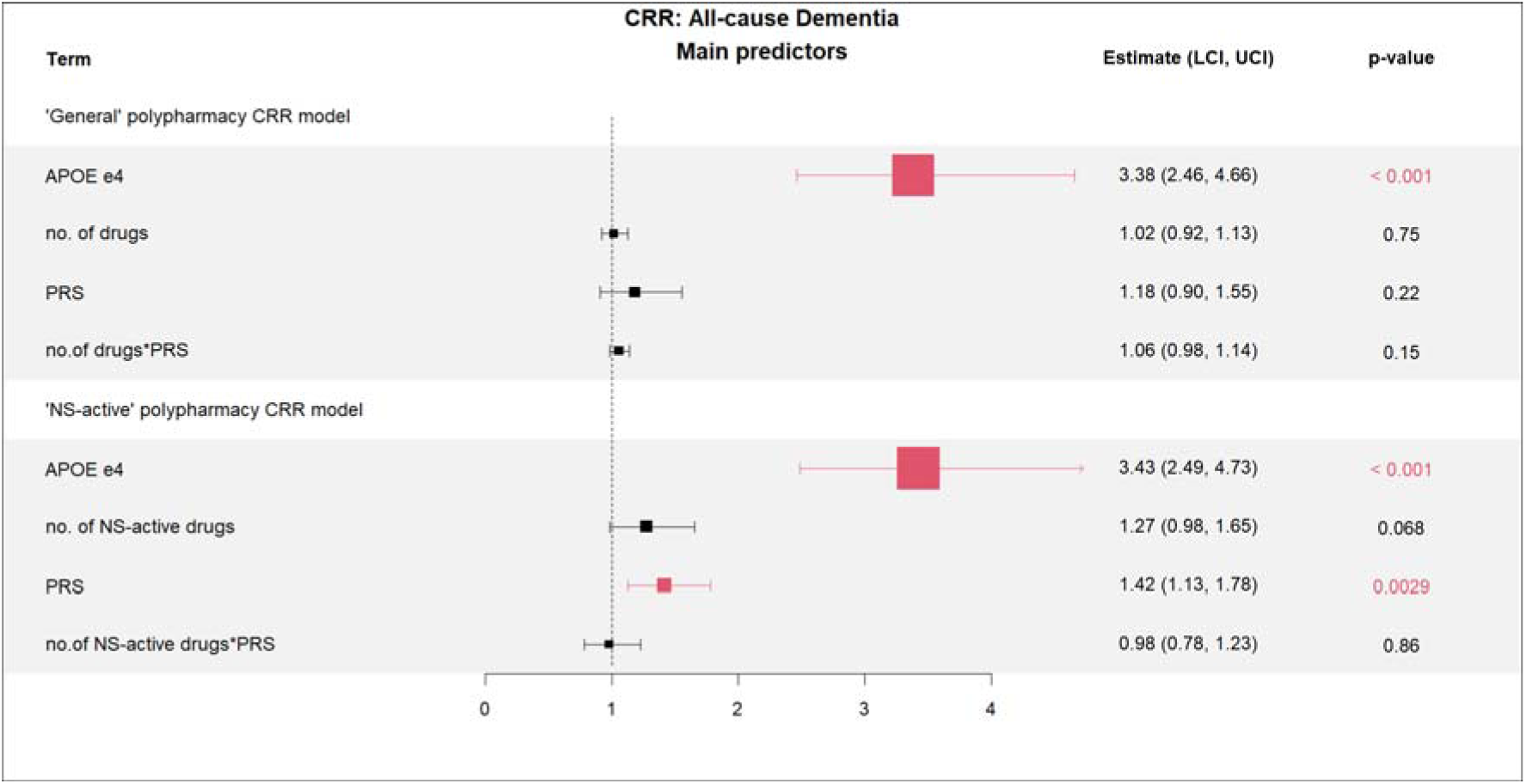
Forest plot showing the Competing Risk Regression models’ Hazard Ratios for the association with all-cause incident dementia, adjusting for death as a competing risk. Only predictors of interest are shown here for illustration purposes, but the full set of predictors were included in the models. Upper figure shows the results for the model using the “general” polypharmacy variable (no. of drugs), whereas lower figure shows the results for the nervous-system active no. of drugs. The “Estimates” of the forest plot are Hazard Ratios (HR). CRR: Competing Risk Regression; UCI: Upper Confidence Interval; LCI: Lower Confidence Interval; PRS: Polygenic Risk Score; NS-active: nervous system-active. The forest plot was produced using the “forest” R package.

Because the interaction term was nonsignificant in both models, the mortality adjusted models were re-run omitting this term. Results showed that PRS had a significant positive effect on dementia risk in both general and NS-active polypharmacy models: HR(general) = 1.39; 95% CI: 1.15-1.67; p<0.001 and HR(NS-active) = 1.40; 95% CI: 1.16-1.69; p-value<0.001 respectively; and that polypharmacy effects were nonsignificant (Sup. Figure 5).

Using AD instead of all-cause dementia as outcome did not meaningfully alter the results of either the “general” or the “nervous system-active” polypharmacy models (Sup. Figure 6 and Sup. Figure 7 for AD-specific Cox PH models and Sup. Figure 8 for AD-specific competing risk regression models).

## 4 DISCUSSION

### 4.1 Interpretation of findings

This is the first study to assess the interaction between polypharmacy and dementia polygenic risk in shaping all-cause dementia risk. Here, higher polypharmacy exposure, higher polygenic risk (PRS), and their interaction were positively associated with increased dementia risk although only PRS was significant when main effects were considered. The positive direction of effect for the interaction between general polypharmacy and PRS is consistent with our hypothesis that the adverse effects of polypharmacy are modulated by adverse genetic effects which increases the risk of dementia over and above their separate effects. However, the nervous system acting polypharmacy interaction with genetic score had an almost zero non-significant effect. It is possible that the low variability of the predictor (only 9 people with 3 or more nervous system active medications) decreased accurate inferences.

Adjusting for death as a competing risk did not substantially change results, indicating that mortality did not affect the association between predictors and dementia. This was unexpected, given the high number of deaths in the sample: 277 people in total and 220 people without dementia that could be potential patients otherwise. In these models, and omitting the nonsignificant polypharmacy x PRS interaction effect, the polygenic effect was significant in the hypothesised direction of increased dementia risk. General and NS-active polypharmacy main effects did not alter the risk of dementia, although the latter effect had a larger HR which approached significance indicating potential differences between types of medication used. In previous studies, PRS and polypharmacy have been often associated with dementia risk^51^, but they had not been modelled together. Our finding of non-significant polypharmacy effects when controlling for AD polygenic risk might indicate that the previously reported polypharmacy effect on dementia is confounded by polygenic risk for the disease. However, this hypothesis may be difficult to prove using PRS. Here low correlation of AD-PRS with polypharmacy was observed (Pearson’s r=0.041), and, overall, polygenic scores for complex diseases explain little variation of drug-related phenotypes^52^. *APOE* status is frequently used as a proxy covariate for genetic susceptibility in studies of polypharmacy effects on dementia^28,53^, with polypharmacy regimes remaining significantly predictive of dementia risk after adjusting for *APOE* status^28^. One study found that polypharmacy lost significance after adjusting for *APOE* in a population of Aboriginal Australians^54^, although the polypharmacy effect size remained similar with its unadjusted value. Larger longitudinal studies of people with both low and high dementia polygenic risk and low and high polypharmacy use are needed to clarify the role of genetic confounding on polypharmacy effects.

The *APOE* gene was the only other covariate that had a significant effect on dementia. This was expected, given its strong and well established role in dementia and AD pathogenesis^8^. Other well established risk factors did not have a significant effect on dementia: BMI, alcohol intake, smoking, and deprivation index, although their effects’ direction was expected^1^. Inadequate sample size is again a plausible explanation, coupled with restrictions of range in the variables stemming from selection bias in the LBC1936: participants are healthier and from higher socioeconomic status than the general population^29^.

In future, polypharmacy x polygenic interaction effects might be investigated with increased pathway specificity. For example, PRS calculated using only SNPs pertaining to a certain biological pathway (e.g. synaptic processes)^12^. Testing whether sub-categories of polypharmacy measurements, such as the nervous system specific polypharmacy predictor, mask the effects of nervous system related pathway-specific PRS could shed light on whether those pathways are modulated by specific drugs. An important caveat, however, is that the predictive ability of pathway-specific PRS are potentially smaller than the genome-wide PRS, so it would be necessary to use large sufficiently powered samples^12^.

Furthermore, when studying polypharmacy, a direct interpretation of associations with outcomes is difficult as confounding by indication may be present^28,55^. This means that dementia may be caused not because of polypharmacy, but due to the comorbidities for which multiple drugs are taken. This can affect both the effects of polypharmacy exposure and the polypharmacy interaction with PRS. However, in the present study, the comorbidities index covariate was not significant and had negligible effect, indicating that the presence of comorbid health conditions did not affect dementia risk. This could be again arising due to small sample size, as per previous similar studies in LBC1936^43^. It could be informative to include separate health conditions that are established dementia risk factors as covariates (e.g., depression, diabetes, and hypertension) and not as part of the comorbidities index covariate. Then, again, small sample size would not allow for the excessive use of covariates, due to the risk of overfitting.

### 4.2 Strengths and limitations

Several strengths can be identified for the present analysis. First, the high accuracy of the dementia ascertainment outcomes, being based on Electronic Health Records, clinical assessments and expert consensus^42^. This increases the power to detect true associations with dementia risk, compared to non-curated dementia diagnoses. Secondly, the LBC1936 study design provides an appropriate dataset to study dementia, due to the older age of the participants (average of 69.5 years on baseline), extensive follow-up period (up to 17.5 years) and absence of dementia at baseline, allowing for the investigation of risk factors early on during disease development. Thirdly, LBC1936’s thorough medication recording protocol, allowing the inclusion of both prescribed and over-the-counter medications in the polypharmacy predictors provides a more accurate picture of the medication state of the individuals. In other studies where polypharmacy is modelled, only prescribed drugs are included^25,28^, which may underestimate the true polypharmacy burden.

On the other hand, several limitations should be acknowledged. The major limitation of the study is the low sample size. Detecting genotype by environment interactions requires very large sample sizes, given their postulated small effect sizes^3^. Additionally, the definition of polypharmacy as the number of (systemic or nervous system acting) drugs taken at a certain time point (Wave 1 testing) is not optimal, as it lacks information on the duration of exposure and adherence to the medication. A better metric would be the number of daily drugs taken^21^ and the time period, however no such information was available. Electronic Health Records data may allow for such modelling. Finally, only baseline (Wave 1) information for all predictors was used (polypharmacy, covariates). Cumulatively incorporating the changes in the predictors over time until dementia diagnosis would improve the power to detect true associations. That is, using information from later waves to construct time-varying predictor variables in the Cox PH models is a sensible future direction for better delineating polypharmacy trajectories.

## 5 CONCLUSIONS

In summary, in the present analysis it was hypothesised that the detrimental effect of polypharmacy on dementia is modulated by one’s genetic profile, with high genetic risk exacerbating adverse polypharmacy effects. This interaction effect was not supported for general polypharmacy or nervous system acting polypharmacy. AD polygenic scores were associated with increased risk of dementia, and there was suggestion that nervous system active drugs had independent polypharmacy effects on dementia risk. Better characterising how polypharmacy exerts adverse effects on individuals with high or low dementia genetic risk may point towards supporting the case for de-prescribing inappropriate medications in older adults dependent on one’s genetic risk towards dementia.

## Data Availability

Information about accessing LBC1936 data can be found here: https://lothian-birth-cohorts.ed.ac.uk/data-access-collaboration

## ACKNOWLEDGEMENTS

We are thankful to Dr. Sarah Harris and Dr. Jure Mur for the insightful discussions on various topics of the project and LBC1936, and to Paul Redmond and Dr. Gail Davies for guidance and assistance on data access.

This research was funded by the Legal & General Group (research grant to establish the independent Advanced Care Research Centre at University of Edinburgh). The funder had no role in conduct of the study, interpretation or the decision to submit for publication. The views expressed are those of the authors and not necessarily those of Legal & General. The LBC1936 study is supported by the Biotechnology and Biological Sciences Research Council (BBSRC) and the ESRC (BB/W008793/1), Age UK (Disconnected Mind project), the Milton Damerel Trust, and the University of Edinburgh. SRC was supported by a Sir Henry Dale Fellowship jointly funded by the Wellcome Trust and the Royal Society (221890/Z/20/Z). The Lothian Birth Cohort 1936 study acknowledges the financial support of NHS Research Scotland (NRS), through Edinburgh Clinical Research Facility. We gratefully acknowledge the contributions of the LBC1936 participants and members of the LBC1936 research team who collect and manage the LBC data.

All authors declare no competing interests.

