## Supplementary Material for "Interplay between polygenic effects and polypharmacy on dementia: An investigation in an elderly Scottish cohort"

**Table of Contents:**

|  | **Page** |
| --- | --- |
| **Supplementary Methods.** Assessment of Cox PH models assumptions. | 3 |
| **Supplementary Table 1.** Dementia subtypes. | 4 |
| **Supplementary Table 2.** Number of individuals taking n number of medications simultaneously at the time of Wave 1 testing, belonging to each anatomical group. | 5 |
| **Supplementary Figure 1.** Plots for each predictor’s Cox PH model Schoenfeld residuals against time (years). | 6 |
| **Supplementary Figure 2.** Martingale Residuals plots of the Cox PH model against the no. of drugs*PRS variable, before and after the removal of an influential individual (top right of the left plot). | 7 |
| **Supplementary Figure 3.** Distribution of participants by polypharmacy and incident dementia status | 8 |
| **Supplementary Figure 4.** Competing Risk Regression models for all-cause incident dementia. | 9 |
| **Supplementary Figure 5.** Competing Risk Regression models for all-cause incident dementia (no interaction). | 10 |
| **Supplementary Figure 6.** Cox PH model for Alzheimer Disease – ‘general’ polypharmacy model. | 11 |
| **Supplementary Figure 7.** Cox PH model for Alzheimer Disease – ‘nervous system-active’ polypharmacy model. | 12 |
| **Supplementary Figure 8.** Competing Risk Regression models for Alzheimer Disease. | 13 |

**Supplementary Methods**

Assessment of Cox PH models assumptions:

First, the proportional hazards assumption was tested, meaning that HRs for each predictor have to be adequately constant at any given time. Graphical and statistical inspection for each predictor did not reveal any predictor that significantly deviated from the proportionality assumption (i.e., no strong association between residuals and time) (Supplementary Figure 1). Subsequently, the assumption for a linear relationship between each predictor and its log-hazard residuals was tested. Here, an influential observation that broke the linearity between the PRS*polypharmacy predictor and the log-hazard was detected and removed (Sup. Figure 2). The assumptions assessments were conducted for both the model including the “general” and the one including the “nervous system-acting” polypharmacy predictor.

**Supplementary Table 1**

Dementia diagnoses by subtype.

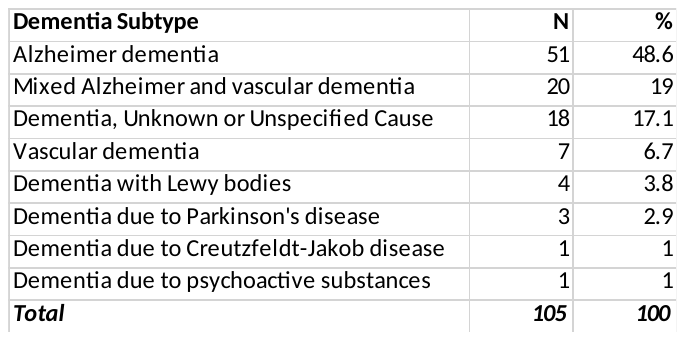

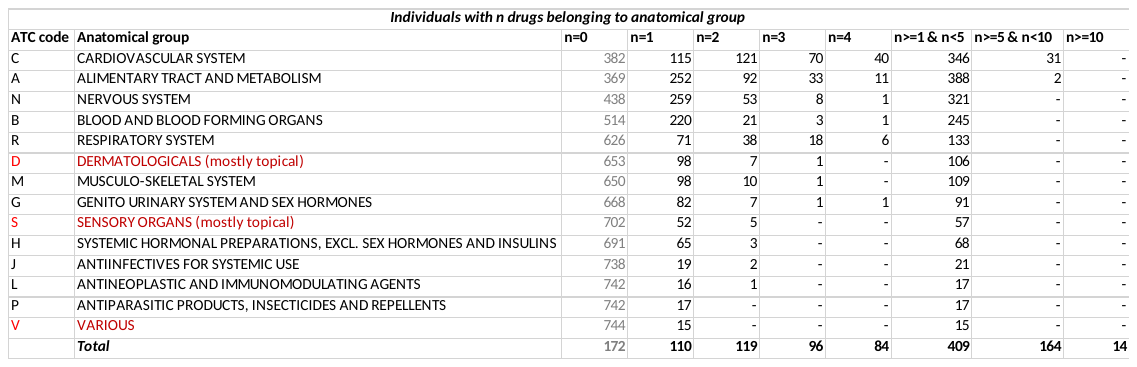
**Supplementary Table 2**

Number of individuals taking n number of medications simultaneously at the time of Wave 1 testing, belonging to each anatomical group. ATC code: corresponds to the 1st level code of the ATC classification system (<https://www.whocc.no/atc_ddd_index/>), denoting each anatomical group defined by the ATC classification system. Anatomical groups in red were excluded from the analysis (see methods section). The “Various” group is defined by the ATC system, containing types of drugs not classified elsewhere. “Total” refers to the any medication taken by individuals, irrespective of ATC code.

**Supplementary Figure 1**

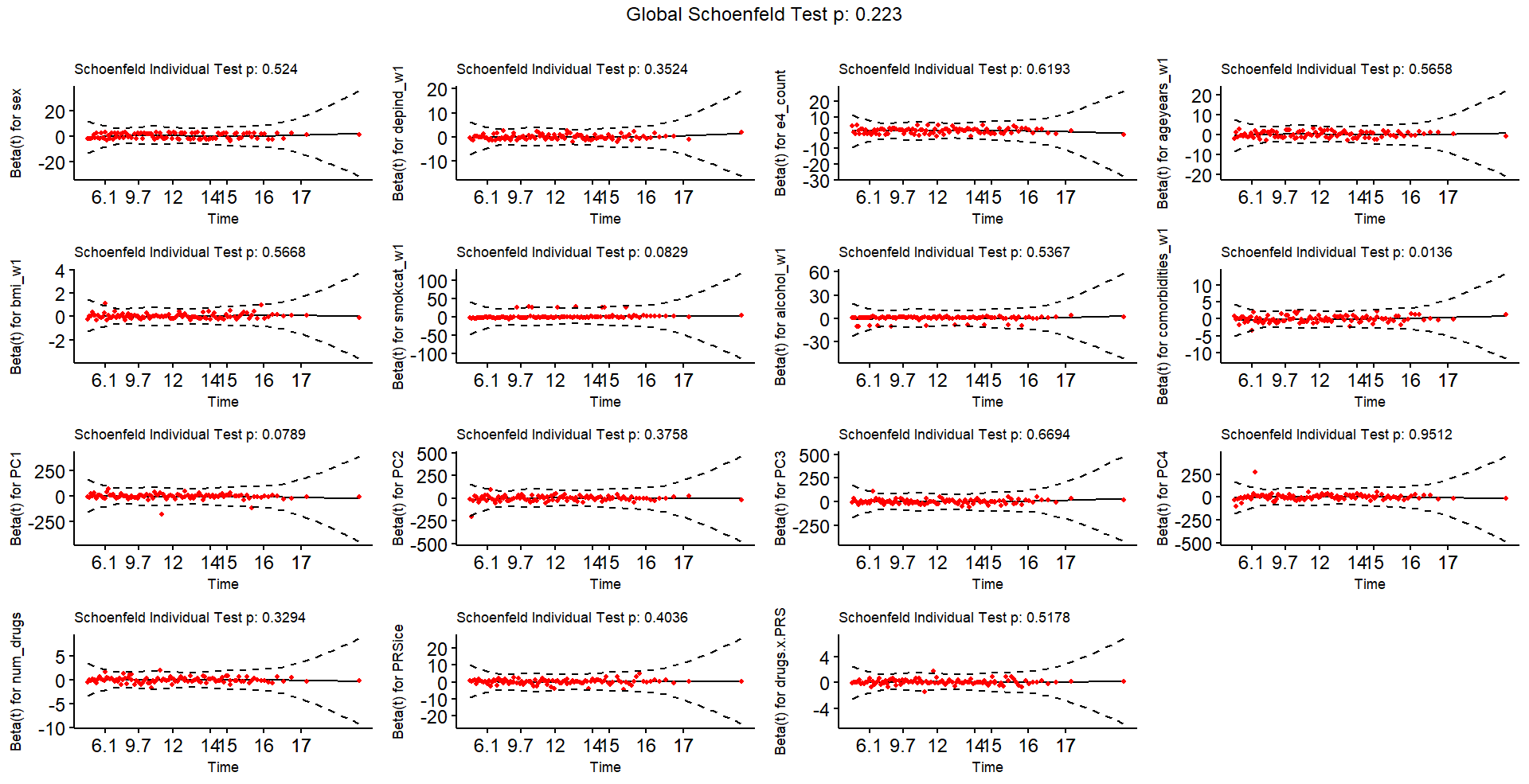

Supplementary Figure 1: Plots for each predictor’s Cox PH model Schoenfeld residuals against time (years). For the proportional hazards assumption to hold, residuals have to be independent of time for each predictor (flat line). Schoenfeld Individual Test performs a formal test for residual-time correlation. Significant p-value would indicate deviation from the PH assumption.

**Supplementary Figure 2**

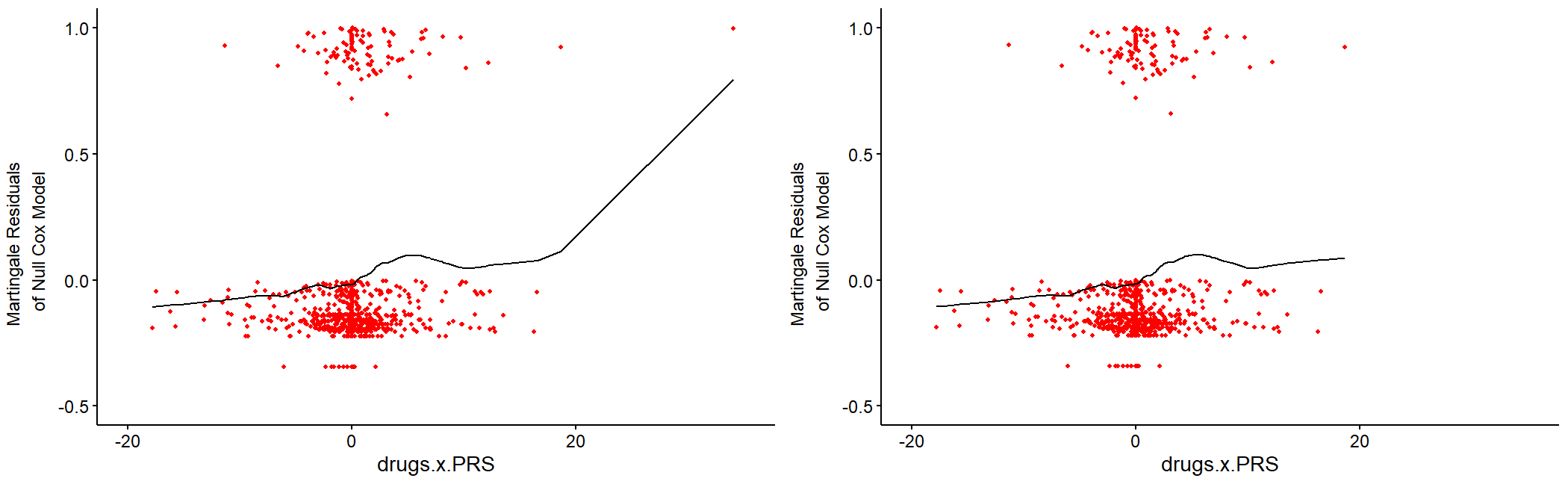

Supplementary Figure 2: Martingale Residuals plots of the Cox PH model against the no. of drugs * PRS variable, before and after the removal of an influential individual (top right of the left plot). The relationship should be adequately linear (straight line) for the linearity assumption to hold. Removing the influential individual produced a more acceptable linear form (right plot).

**Supplementary Figure 3**

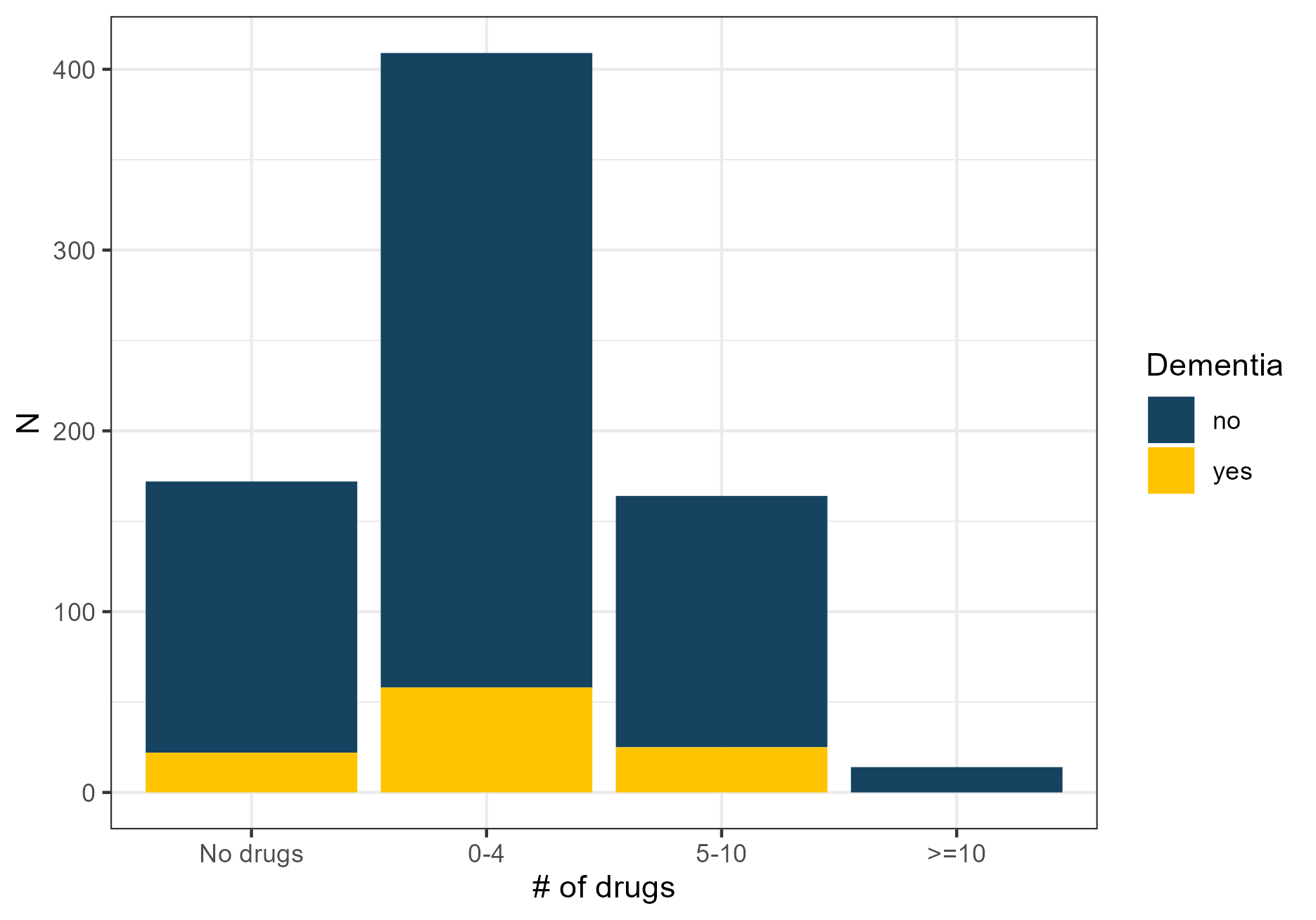

Supplementary Figure 3: Distribution of participants by polypharmacy and incident dementia status. N: number of individuals according to drugs taken at Wave 1 and dementia diagnosis at follow-up.

**Supplementary Figure 4**

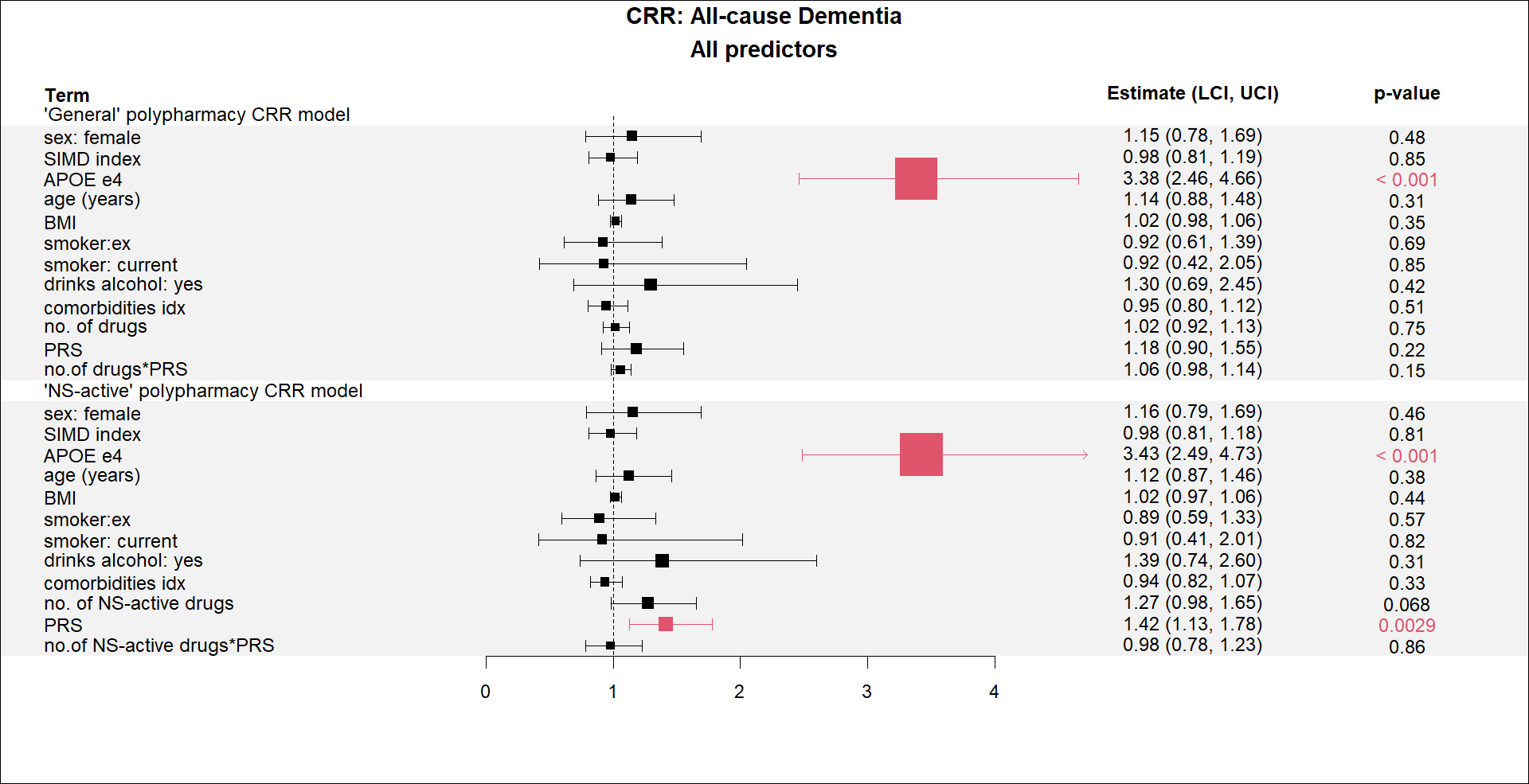

Supplementary Figure 4: Competing Risk Regression models for all-cause incident dementia. The full set of predictors are shown here. Part of this forest plot is show in Figure 3 of the Results section. Upper figure shows the results for the model using the “general” polypharmacy variable (no. of drugs), whereas lower figure shows the results for the nervous-system active no. of drugs. The “Estimates” of the forest plot are Hazard Ratios (HR). CRR: Competing Risk Regression; UCI: Upper Confidence Interval; LCI: Lower Confidence Interval; PRS: Polygenic Risk Score; NS-active: nervous system-active BMI: Body Mass Index; SIMD: Scottish Index of Multiple Deprivation. The forest plot was produced using the “forest” R package.

**Supplementary Figure 5**

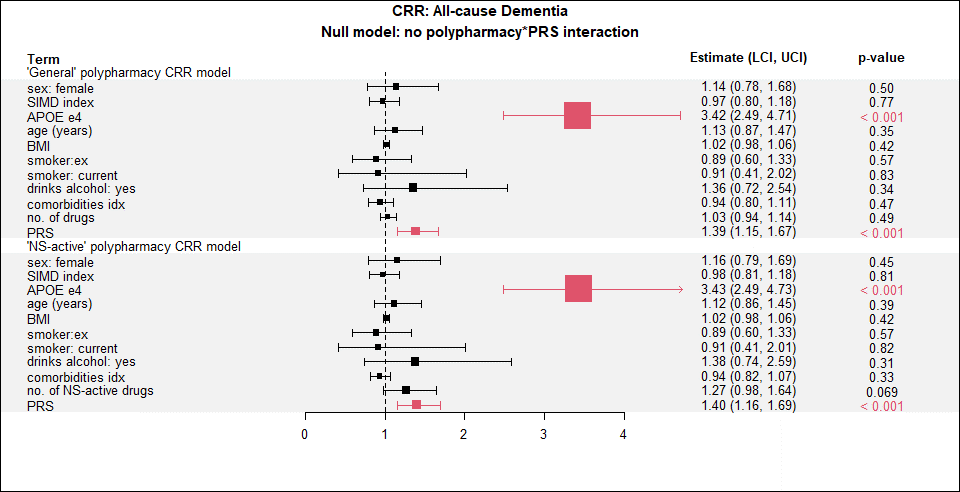

Supplementary Figure 5: Competing Risk Regression models for all-cause incident dementia (no interaction). The full set of predictors are shown here for the set of null models that did not include the interaction term between polypharmacy and PRS. Upper figure shows the results for the model using the “general” polypharmacy variable (no. of drugs), whereas lower figure shows the results for the nervous-system active no. of drugs. The “Estimates” of the forest plot are Hazard Ratios (HR). CRR: Competing Risk Regression; UCI: Upper Confidence Interval; LCI: Lower Confidence Interval; PRS: Polygenic Risk Score; NS-active: nervous system-active BMI: Body Mass Index; SIMD: Scottish Index of Multiple Deprivation. The forest plot was produced using the “forest” R package.

**Supplementary Figure 6**

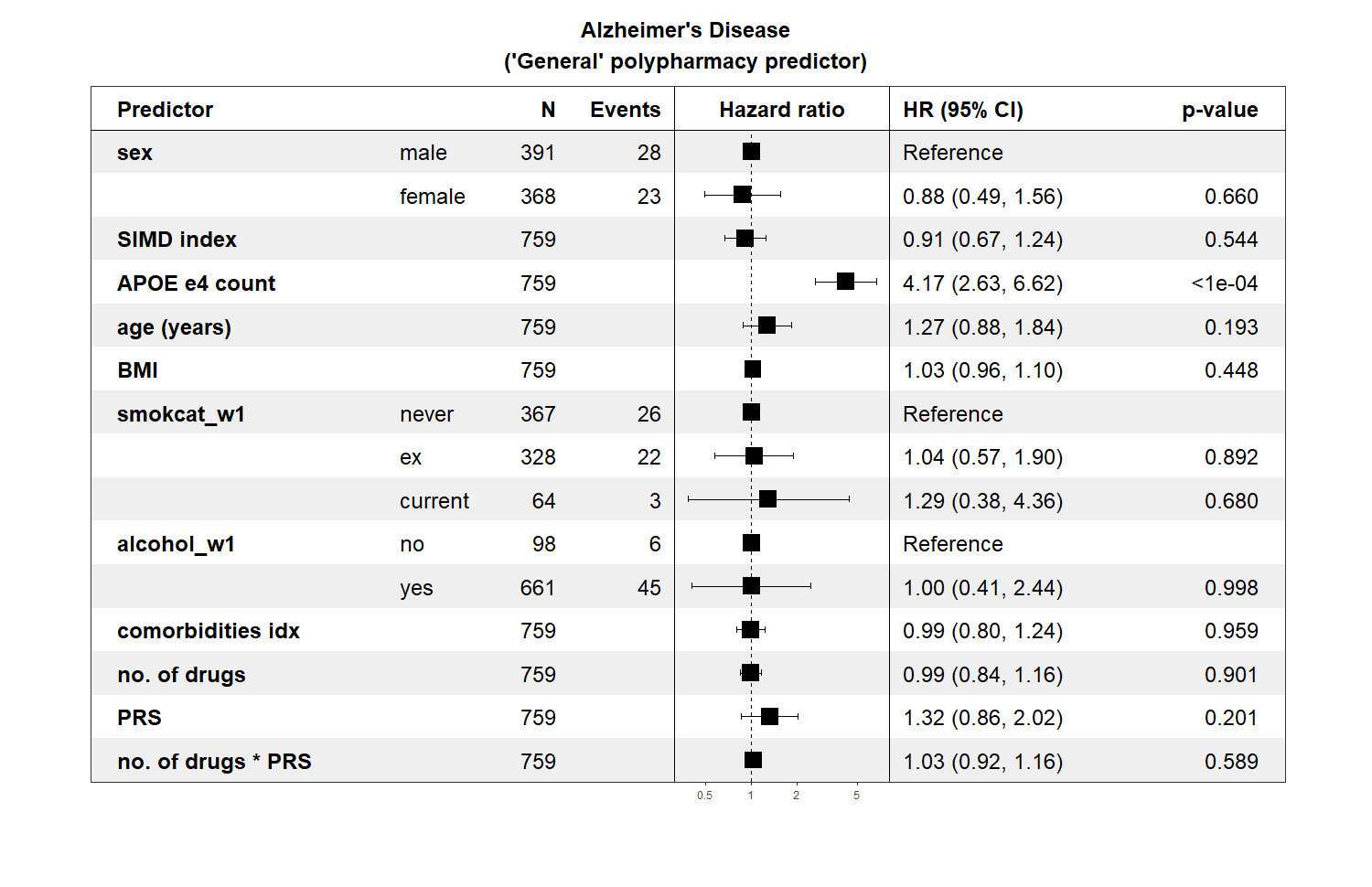

Supplementary Figure 6: Cox PH model for Alzheimer Disease – ‘general’ polypharmacy model. The “no. of drugs” predictor includes all drugs taken by an individual after data cleaning. HR for Genomic Principal Components are not shown for illustration purposes, but were included in the model. The forest plot was produced using the “forestmodel” R package. HR: Hazard Ratio; CI: Confidence Interval; PRS: Polygenic Risk Score; BMI: Body Mass Index; SIMD: Scottish Index of Multiple Deprivation.

**Supplementary Figure 7**

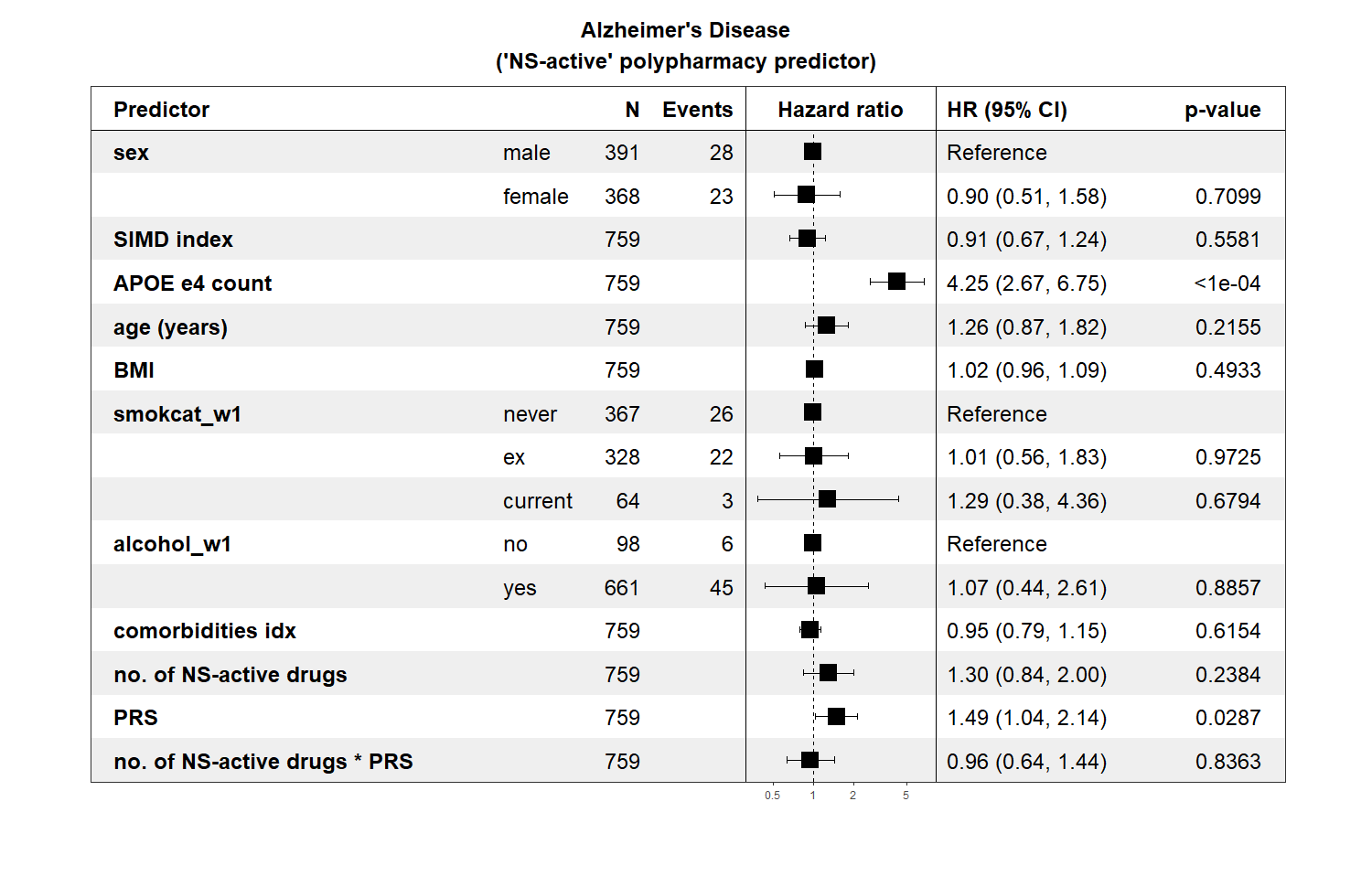

Supplementary Figure 7: Cox PH model for Alzheimer Disease – ‘nervous system-active’ polypharmacy model. The “no. of NS-active drugs” predictor includes drugs acting on the nervous system taken by an individual. HR for Genomic Principal Components are not shown for illustration purposes, but were included in the model. The forest plot was produced using the “forestmodel” R package. HR: Hazard Ratio; CI: Confidence Interval; PRS: Polygenic Risk Score; BMI: Body Mass Index; SIMD: Scottish Index of Multiple Deprivation; NS-active: nervous system-active.

**Supplementary Figure 8**

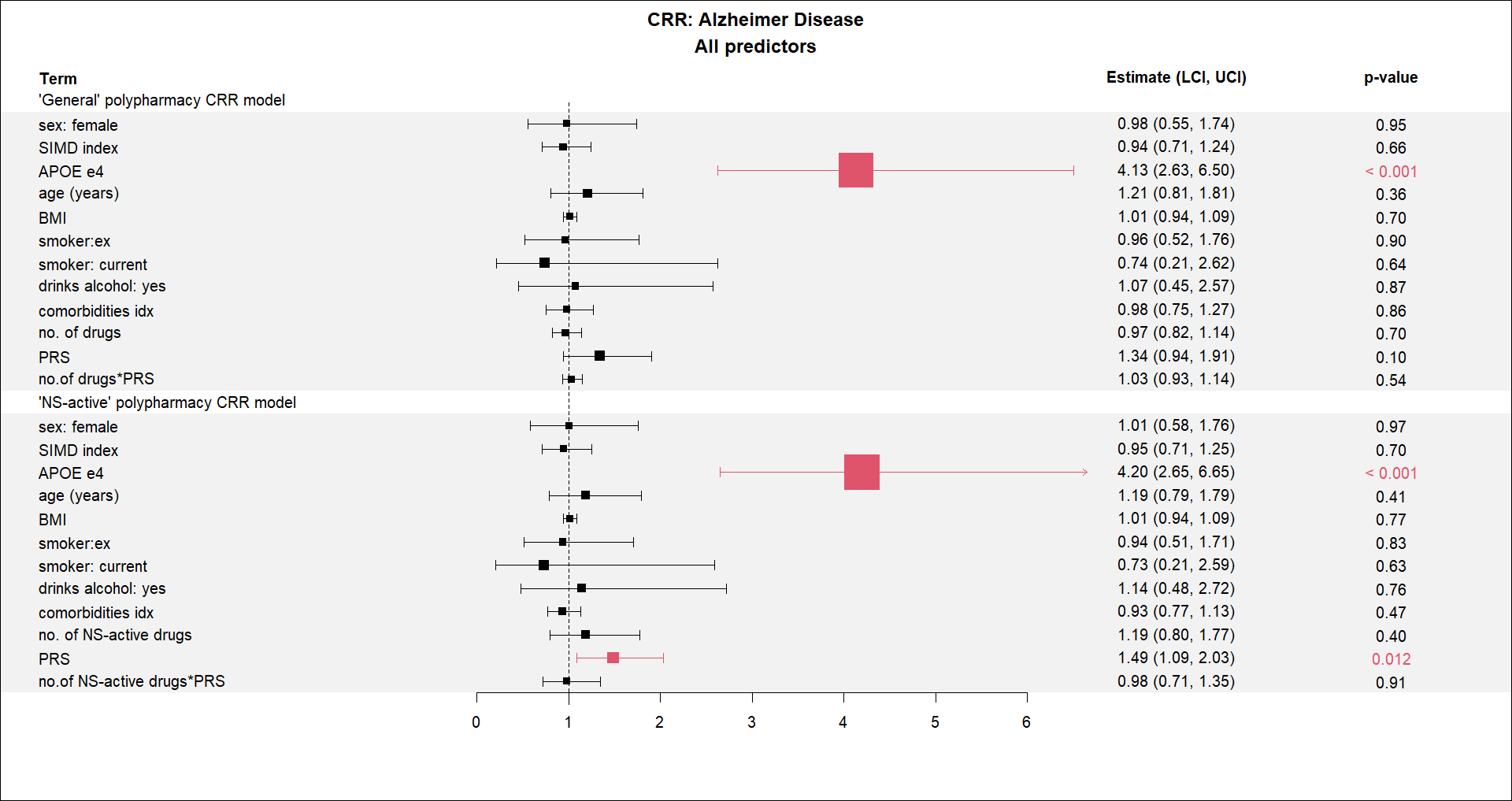

Supplementary Figure 8: Competing Risk Regression models for Alzheimer Disease. The full set of predictors are shown here. Upper figure shows the results for the model using the “general” polypharmacy variable (no. of drugs), whereas lower figure shows the results for the nervous-system active no. of drugs. The “Estimates” of the forest plot are Hazard Ratios (HR). CRR: Competing Risk Regression; UCI: Upper Confidence Interval; LCI: Lower Confidence Interval; PRS: Polygenic Risk Score; NS-active: nervous system-active BMI: Body Mass Index; SIMD: Scottish Index of Multiple Deprivation. The forest plot was produced using the “forest” R package.
